# Association Between Disability and Longitudinal Brain Volumetric Measures Across the Peri-Pregnancy Period in Women with Multiple Sclerosis: A Pilot Study

**DOI:** 10.64898/2026.09.06.26362381

**Authors:** Meryem Tuba Sönmez, Sümeyye Koç, Korhan Buyukturkoglu, Levi J. Davis, Duygu Arslan Mehdiyev, Murat Terzi, Ceren Tozlu

**Author notes:** **Corresponding Author:** Meryem Tuba Sönmez, **Mailing address:** Ankara Etlik City Hospital, Neurology Clinic, Yenimahalle, Ankara, Turkiye, **Email:**.

## Abstract

**Background:** Pregnancy is a distinct neuroendocrine and immunological state in women with multiple sclerosis (MS). Relapse activity typically decreases during gestation and increases after delivery, but the relationship between peri-pregnancy brain volumetric measures and disability remains unclear.

**Methods:** This retrospective longitudinal pilot study included 14 women with relapsing-remitting MS who underwent MRI examinations before pregnancy and after delivery. Thirty-eight MRI scans were analyzed, with a mean of 2.71 ± 0.99 timepoints per patient. Global and regional brain volumes were measured using the longitudinal FreeSurfer pipeline. Choroid plexus (CP) volume was evaluated as an exploratory neuroimmune imaging marker. Associations between volumetric measures and Expanded Disability Status Scale (EDSS) scores were assessed using linear mixed-effects modeling with forward stepwise selection.

**Results:** The final EDSS model retained bilateral ventral diencephalon (ventral DC) volumes, left amygdala volume, right pallidum volume, left nucleus accumbens volume, and cerebral cortex volume. The most prominent association involved the left ventral DC, where lower volume was associated with higher EDSS scores (p = 0.0008). CP volume showed no significant longitudinal change across peri-pregnancy MRI timepoints (p = 0.21) and was not retained in the final EDSS model.

**Conclusion:** Regional brain volumetric measures, particularly ventral DC volumes, were associated with disability across the peri-pregnancy period despite low EDSS levels. To our knowledge, this is the first longitudinal evaluation of CP volume across the peri-pregnancy period in women with MS; no significant change was observed over time. These exploratory findings require confirmation in larger prospective studies using standardized serial MRI protocols.

## Introduction

Multiple sclerosis (MS) predominantly affects women of reproductive age, making pregnancy an important clinical period in the disease course [1]. Pregnancy induces marked hormonal and immunological adaptations [2] that are accompanied by reduced relapse activity, particularly during the third trimester, followed by a transient increase in relapse risk after delivery [3–6]. Although these inflammatory patterns are well established, the accompanying structural changes in the brain are less well understood.

Recent longitudinal MRI studies in healthy women have demonstrated dynamic structural brain changes during pregnancy and postpartum, including reductions in cortical and subcortical gray matter volumes, ventricular enlargement, and changes in white matter microstructure [7, 8]. Some of these changes show partial recovery during the postpartum period [8, 9]. In women with MS, evidence on peri-pregnancy structural brain changes remains limited, with previous studies reporting thalamic volume reduction and accelerated brain volume loss after pregnancy [10, 11]. Regional brain volumes, particularly those of deep gray matter structures, have been linked to disability in MS [12]. Pregnancy-related physiological remodeling in women with MS may therefore overlap with disease-related structural changes. How these regional changes relate to disability during this period, however, remains unclear.

The choroid plexus (CP), an interface between peripheral immunity and the central nervous system, has gained attention as a potential imaging marker in MS [13]. Larger CP volume has been associated with inflammatory activity, gray matter loss, thalamic atrophy, and disability, but its relevance during the peri-pregnancy period is unknown [14–17].

Based on this evidence, we hypothesized that regional brain volumetric measures would be associated with disability during the peri-pregnancy period. In this longitudinal pilot MRI study, we examined global and regional brain volumes in women with relapsing-remitting MS before pregnancy and after delivery and assessed their associations with disability. We also evaluated CP volume as an exploratory neuroimmune imaging marker.

## Methods

### Study Design and Participants

This retrospective longitudinal study was conducted at the Department of Neurology, Ondokuz Mayıs University Hospital. The study included women who had given birth and were diagnosed with relapsing-remitting multiple sclerosis (RRMS) according to the 2017 McDonald criteria [18]. Patients were identified through the institutional MS registry and electronic medical records.

Eligible patients were required to have at least one pre-pregnancy brain MRI obtained within the two years before delivery and at least one postpartum brain MRI obtained within the two years after delivery. This peri-pregnancy window was selected to keep MRI examinations temporally close to pregnancy and to minimize the potential contribution of aging and disease duration to brain volume loss.

Given the limited number of patients with eligible longitudinal peri-pregnancy MRI data, two scans with minor deviations from the predefined window were retained: one pre-pregnancy scan obtained 24.1 months before delivery and one postpartum scan obtained 25.2 months after delivery, each from a different patient.

### Clinical Data Collection

Demographic, clinical, and MRI data were obtained from medical records and the institutional MS registry. Missing or outdated clinical and peri-pregnancy information was supplemented through individual patient interviews. All MRI scans and longitudinal study data were reviewed by the investigators to ensure completeness and consistency across study timepoints. Disability was assessed using the Expanded Disability Status Scale (EDSS), scored by the treating neurologist at the clinical visit closest to each MRI examination.

The following variables were recorded: age at delivery, dates of MS symptom onset and diagnosis, disease duration from symptom onset to delivery, pre-pregnancy disease-modifying therapy (DMT) use and timing of discontinuation, relapse history in the two years before conception, relapses during pregnancy and in the postpartum period, delivery mode, and breastfeeding status and duration.

DMTs were categorized by treatment efficacy as low-efficacy/platform therapies (interferon beta-1a, pegylated interferon beta-1a, glatiramer acetate), moderate-efficacy therapy (dimethyl fumarate), and high-efficacy therapies (natalizumab and fingolimod).

Before analysis, all clinical and MRI data were anonymized, and each participant was assigned a study-specific identification code.

### MRI Acquisition and Processing

All MRI examinations were performed as part of routine clinical care on 1.5-T or 3-T scanners. Scanner manufacturers included Philips, Siemens, GE, and Hitachi. T1-weighted and fluid-attenuated inversion recovery (FLAIR) sequences were used in this study. Details of the scanners and acquisition parameters are provided in the Supplementary Table.

As a first step in the MRI analysis, the N4 bias correction algorithm was applied to reduce low-frequency intensity nonuniformities in the images [19]. Considering the potential effects of T1-hypointense lesions on tissue segmentation [20, 21], lesion-inpainted images were used for region of interest (ROI) mask generation. Automated lesion segmentation was performed using SAMSEG [22], a module within FreeSurfer, followed by lesion inpainting using FSL (FMRIB Software Library).

FreeSurfer version 7.4.1 was subsequently applied to lesion-inpainted T1-weighted images using the longitudinal processing stream to obtain global and regional volumetric measures across timepoints [23]. This framework improves reliability and statistical power by generating an unbiased within-subject template from all available timepoints and using this template to initialize subsequent processing steps, thereby reducing variability and increasing sensitivity to subtle longitudinal changes in brain structure.

Because of the small size, irregular morphology, and relatively low tissue contrast of the CP on conventional MRI, FreeSurfer-derived CP segmentations were not used directly. Previous studies have demonstrated limited accuracy of automated FreeSurfer segmentation for this structure [24]. The initial CP segmentation was obtained using FreeSurfer and was subsequently refined using a Gaussian Mixture Model (GMM)-based segmentation algorithm, which improves tissue classification by modeling voxel intensity distributions within anatomically constrained regions [24]. The resulting masks were visually inspected and manually corrected when necessary to ensure anatomical accuracy. Final CP volumes derived from these refined segmentations were used in all subsequent analyses.

The neuroimaging predictors included bilateral FreeSurfer-derived volumes of the thalamus, caudate nucleus, putamen, pallidum, hippocampus, amygdala, nucleus accumbens, and ventral diencephalon (ventral DC). Global brain volumetric measures included cerebral white matter volume, cerebral gray matter volume, cerebral cortex volume, cerebellar white matter volume, and cerebellar gray matter volume. CP volume was also included.

Inter-scanner variability was addressed using longitudinal ComBat, which adjusted for scanner manufacturer (Siemens, Philips, GE, or Hitachi), field strength, and acquisition type (2D vs 3D), with age and estimated total intracranial volume included as covariates [25].

### Statistical Analysis

Continuous variables were summarized using mean ± standard deviation or median and interquartile range, as appropriate based on their distribution. Categorical variables were summarized as counts and percentages.

Given the limited sample size and the large number of candidate MRI-derived volumetric predictors, forward stepwise variable selection was used to obtain a parsimonious linear mixed-effects model for EDSS. Candidate predictors were sequentially evaluated, and those that significantly improved model fit were retained in the final model. The candidate predictors were the harmonized volumetric measures, including cerebral white matter volume, cerebral gray matter volume, cerebral cortex volume, cerebellar white matter volume, cerebellar gray matter volume, CP volume, and bilateral volumes of the thalamus, caudate nucleus, putamen, pallidum, hippocampus, amygdala, nucleus accumbens, and ventral DC. EDSS was used as the outcome variable. A random intercept for each participant was included to account for repeated measures within individuals. Following forward stepwise selection, session number was included as a fixed effect representing MRI timepoint in the final linear mixed-effects model.

In addition, longitudinal change in CP volume across peri-pregnancy MRI timepoints was assessed using a separate linear mixed-effects model, with a random intercept for each participant. Statistical significance was set at p < 0.05. All analyses were conducted in R version 4.4.3.

## Results

### Study Cohort and Pregnancy-Related Characteristics

The study included 14 women with relapsing-remitting multiple sclerosis who had given birth and had longitudinal peri-pregnancy MRI data available. The mean age at delivery was 28 ± 5 years, and the mean age at MS symptom onset was 22.4 ± 2.6 years. The median disease duration from symptom onset to delivery was 6 years. Pre-pregnancy disability was low, with a median EDSS score of 0. Demographic, clinical, and pregnancy-related characteristics are summarized in Table 1.

**Table 1.** Demographic, Clinical, and Pregnancy Characteristics of the Study Cohort.

| Characteristic | All Participants (n = 14) |
| --- | --- |
| **A. Demographic and Clinical Characteristics** |  |
| Age at MS symptom onset, years, mean ± SD [range] | 22.4 ± 2.6 [19–28] |
| Disease duration from symptom onset to delivery, years, median (IQR) [range] | 6.0 (2.4–10.3) [0.9–16.2] |
| MS subtype, RRMS, n (%) | 14 (100) |
| OCB status |  |
| Type 1, n (%) | 1 (7.1) |
| Type 2, n (%) | 9 (64.3) |
| Not performed, n (%) | 4 (28.6) |
| Age at delivery, years, mean ± SD [range] | 28.0 ± 5.0 [20–37] |
| EDSS at pre-pregnancy MRI, median (IQR) [max] | 0.0 (0.0–1.0) [2.0] |
| **B. Pregnancy and Treatment** |  |
| Pre-pregnancy DMT, n (%) | 12 (85.7) |
| Platform/low-efficacy (IFN β-1a, pegIFN β-1a, glatiramer acetate), n (%) | 6 (42.9) |
| Moderate-efficacy (dimethyl fumarate), n (%) | 2 (14.3) |
| High-efficacy (natalizumab, fingolimod), n (%) | 4 (28.6) |
| No DMT, n (%) | 2 (14.3) |
| DMT discontinuation timing |  |
| Upon pregnancy confirmation, n (%) | 5 (35.7) |
| of whom high-efficacy DMT, n | 3 |
| Planned preconception, n (%) | 7 (50.0) |
| Pre-pregnancy relapse ($\geq 1$ in prior 2 years), n (%) | 7 (50.0) |
| Relapse during pregnancy, n (%) | 1 (7.1) |
| Postpartum relapse, n (%) | 6 (42.9) |
| Delivery mode |  |
| Cesarean section, n (%) | 9 (64.3) |
| Vaginal, n (%) | 5 (35.7) |
| Breastfeeding, n (%) | 14 (100) |
| Breastfeeding duration, months, median (IQR) [range] | 8 (4–16) [2–24]<sup>a</sup> |

Abbreviations: DMT, disease-modifying therapy; EDSS, Expanded Disability Status Scale; IFN, interferon; IQR, interquartile range; MS, multiple sclerosis; OCB, oligoclonal bands; pegIFN, pegylated interferon; RRMS, relapsing-remitting multiple sclerosis; SD, standard deviation.
<sup>a</sup> Five patients were still breastfeeding at the time of data collection; reported durations represent minimum breastfeeding periods for these patients.

Before pregnancy, 12 patients were receiving disease-modifying therapy, including low-efficacy/platform therapies in six patients, dimethyl fumarate in two patients, and high-efficacy therapies in four patients. DMT was discontinued after pregnancy confirmation in five patients and as part of planned preconception management in seven patients. Seven patients had experienced at least one relapse during the two years before conception. One patient experienced a relapse during pregnancy, and six experienced a postpartum relapse. All participants breastfed, with a median breastfeeding duration of 8 months.

### MRI Dataset

A total of 38 MRI scans were analyzed, with a mean of 2.71 ± 0.99 timepoints per patient. The median interval between the pre-pregnancy MRI and delivery was 13.5 months. The median intervals from delivery to the first and last postpartum MRI scans were 3.7 and 10.9 months, respectively. MRI acquisition characteristics are presented in Table 2.

**Table 2.** MRI acquisition characteristics.

| Characteristic | Value |
| --- | --- |
| Total MRI scans analyzed, n | 38 |
| MRI timepoints per patient, mean ± SD [range] | 2.71 ± 0.99 [2–5] |
| Pre-pregnancy MRI to delivery, months, median (IQR) [range] | 13.5 (11.9–19.5) [8.5–24.1]<sup>a</sup> |
| Delivery to first postpartum MRI, months, median (IQR) [range] | 3.7 (2.0–10.9) [1.1–25.2]<sup>b</sup> |
| Delivery to last postpartum MRI, months, median (IQR) [range] | 10.9 (7.0–15.2) [1.9–25.2] |
| **MRI field strength** |  |
| 1.5 Tesla | 24 |
| 3.0 Tesla | 14 |
| **Scanner manufacturer, n** |  |
| Philips / Siemens / GE / Hitachi | 21 / 9 / 7 / 1 |

Abbreviations: IQR, interquartile range; SD, standard deviation.
<sup>a</sup> One patient had a pre-pregnancy MRI obtained 24.1 months before delivery; given the minor deviation and the retrospective real-world nature of the study, this scan was retained in the analysis.
<sup>b</sup> One patient had the first postpartum MRI at 25.2 months after delivery; excluding this outlier, the median interval was 3.2 months (IQR 1.9–7.3).

### Longitudinal Disability Course

Among the 14 women with MS, the mean EDSS was 0.57 at baseline and 0.60 at the last follow-up. The mean within-patient change in EDSS between baseline and the last follow-up was 0.035, indicating only a small average change in EDSS over follow-up.

### Association Between Brain Volumetric Measures and EDSS

Linear mixed-effects modeling was used to evaluate the longitudinal association between MRI-derived volumetric measures and EDSS across peri-pregnancy timepoints. Forward stepwise selection retained left and right ventral DC volumes, left amygdala volume, right pallidum volume, left nucleus accumbens volume, and cerebral cortex volume. Session number was subsequently included as a fixed effect representing MRI timepoint in the final EDSS model. The most prominent regional association involved the left ventral DC, where lower volume was associated with higher EDSS scores (p = 0.0008). Model estimates and p values are presented in Table 3. Individual longitudinal trajectories of left and right ventral DC volumes are shown in Figure 1, illustrating considerable inter-individual variability across the peri-pregnancy period.

**Table 3.** Results of the Linear Mixed-Effects Model for EDSS During the Peri-Pregnancy Period.

| Predictor | Estimate ($\beta$) | P value |
| --- | --- | --- |
| Session (timepoint) | 2.934e-01 | 0.002 |
| Left amygdala volume | 3.031e-03 | 0.025 |
| Left ventral diencephalon volume | -2.637e-03 | 0.0008 |
| Right ventral diencephalon volume | 4.112e-03 | 0.001 |
| Right pallidum volume | -2.050e-03 | 0.028 |
| Left nucleus accumbens volume | -4.349e-03 | 0.005 |
| Cerebral cortex volume | 1.459e-05 | 0.03 |

Abbreviations: EDSS, Expanded Disability Status Scale.
Candidate volumetric predictors were evaluated using forward stepwise selection. Session number was subsequently included as a fixed effect representing MRI timepoint in the final mixed-effects model. Only variables included in the final model are shown.

**Fig. 1.**
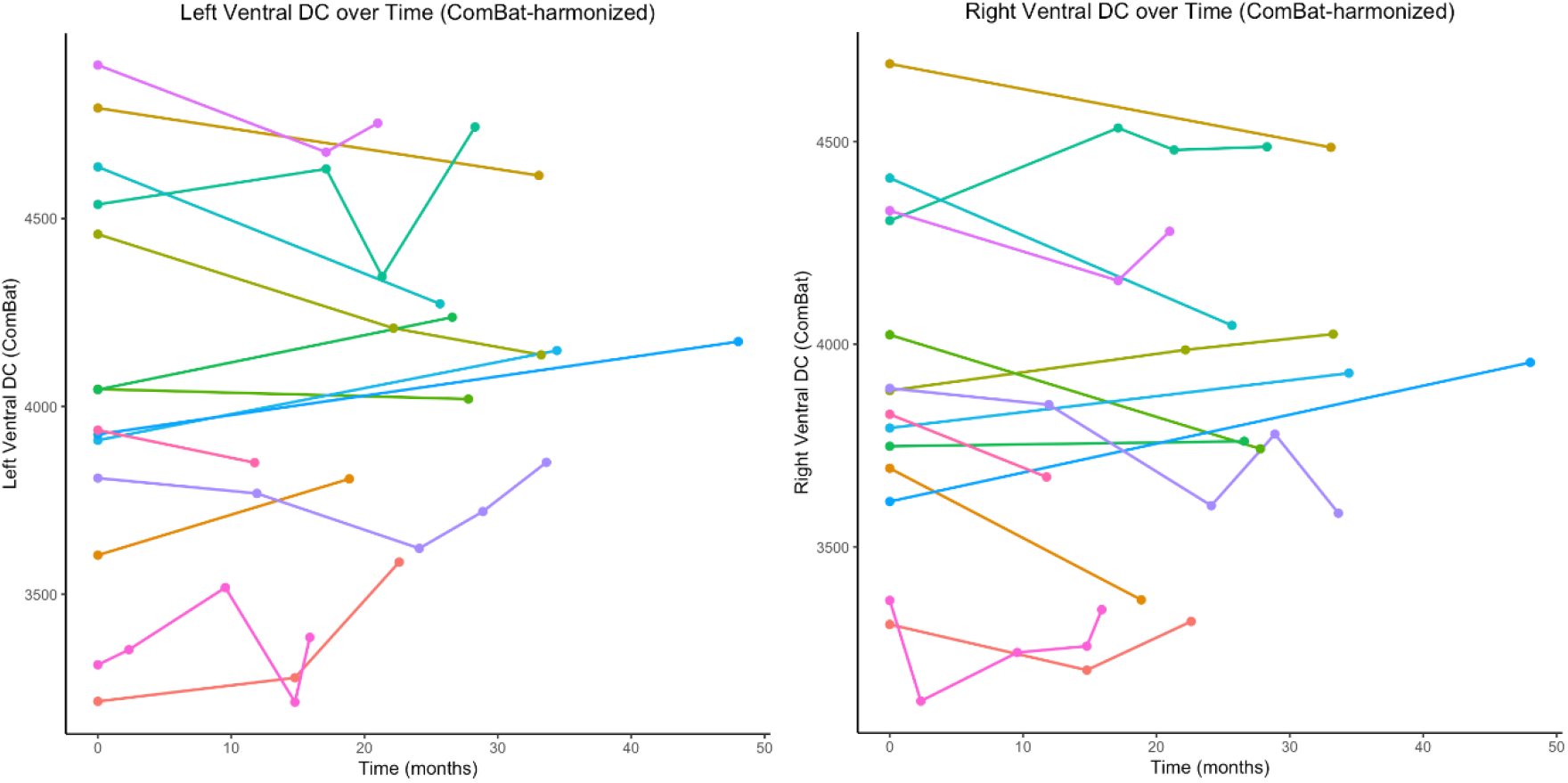
Individual longitudinal trajectories of left and right ventral diencephalon volumes across the peri-pregnancy period. Each line represents a single participant across available MRI timepoints. Volumes are shown after harmonization and preprocessing as described in the methods.

### Choroid Plexus Volume

In the forward stepwise mixed-effects model evaluating MRI-derived predictors of EDSS, CP volume was not retained in the final model. In a separate linear mixed-effects model evaluating longitudinal change in CP volume across peri-pregnancy MRI timepoints, no significant change was observed (p = 0.21). Individual trajectories of CP volume are shown in Supplementary Figure.

## Discussion

Our findings partially supported our hypothesis that regional brain volumetric measures would be associated with disability during the peri-pregnancy period. The most prominent association involved the left ventral DC, where lower volume was associated with higher EDSS scores. Additional associations were observed for the right ventral DC, left amygdala, right pallidum, left nucleus accumbens, and cerebral cortex. CP volume showed no significant longitudinal change and was not retained in the final EDSS model.

Previous MRI studies examining pregnancy-related structural brain changes in women with MS have reported heterogeneous findings. A preliminary study found thalamic volume reduction even in clinically stable patients [10], while Uher et al. [11] reported increased lesion burden and accelerated brain volume loss after pregnancy despite relatively stable disability. In contrast, Lorefice et al. [26] found no association between pregnancy history and long-term whole-brain, white matter, or gray matter volumes in a larger real-world cohort. A recent review integrating evidence from healthy pregnancy and MS similarly concluded that the structural effects of pregnancy on the MS brain remain poorly understood [27]. Indeed, most previous studies have focused on global measures or individual regions, leaving it unclear how multiple regional volumes relate to disability across the peri-pregnancy period. Our study addresses this gap.

The ventral DC is a composite subcortical region that includes the hypothalamus, mammillary bodies, substantia nigra, red nucleus, and lateral geniculate nucleus [8]. Its hypothalamic component is especially noteworthy given the marked neuroendocrine adaptations of gestation. Placental signaling alters hypothalamic–pituitary–adrenal function, while circulating estrogen, progesterone, and glucocorticoid levels rise across pregnancy and fall rapidly after delivery [28]. In MS, this hormonal transition is accompanied by immunological changes that have been linked to the characteristic clinical course across pregnancy and postpartum [2]. In a longitudinal precision neuroimaging study of a single healthy pregnancy, bilateral ventral DC volumes decreased across gestation, with the right ventral DC showing the strongest negative association with gestational progression and incomplete postpartum recovery [8]. Outside the pregnancy setting, longitudinal data from the FutureMS cohort demonstrated bilateral ventral DC volume loss over one year in recently diagnosed RRMS [29]. Fujimori et al. [30] further reported that ventral DC volume differed across patient clusters characterized by increasing levels of disability. Against this background, our finding that lower left ventral DC volume was associated with higher EDSS suggests that this region may have particular clinical relevance during the peri-pregnancy period in women with MS. However, the present data cannot determine the relative contributions of MS-related structural change and pregnancy-related physiological remodeling to this association.

Other predictors retained in the final model included the volumes of the left amygdala, right pallidum, left nucleus accumbens, and cerebral cortex. These structures are involved in motor, cognitive, emotional, and reward-related processes. Changes in amygdala, pallidal, and nucleus accumbens volumes have been associated with changes in EDSS, although the direction and temporal pattern of these associations varied across regions [31]. In other MS cohorts, cortical and subcortical gray matter volumes have likewise been associated with disability [32–34]. At the same time, cortical and subcortical brain regions have been shown to undergo dynamic structural changes during pregnancy and postpartum in healthy women [7–9]. Therefore, the regional associations observed in our peri-pregnancy MS cohort should be considered alongside MS-related structural changes and normal pregnancy-related brain adaptations. Because the EDSS is weighted toward physical and ambulatory disability, it may not fully capture the broader clinical relevance of these regional findings.

The direction of the associations varied across the regional predictors retained in the final model. Most notably, left and right ventral DC volumes showed significant associations with EDSS in opposite directions. The basis of this hemispheric asymmetry remains uncertain, and a true lateralized biological effect cannot be established from the present data. One possible explanation is model instability related to the combination of correlated regional volumetric measures and the limited sample size. Similar methodological concerns have been reported in MS volumetric studies, where correlated MRI features can complicate feature selection and contribute to unstable model outputs [35]. The direction and hemispheric pattern of these associations should therefore be interpreted cautiously and confirmed in larger longitudinal cohorts.

We examined CP volume because of its proposed role as an imaging biomarker of neuroinflammation. A recent meta-analysis showed that normalized CP volume is increased in people with MS, while its association with EDSS was modest and the strongest association was with ventricular volume [17]. In addition, the CP contains estrogen, progesterone, and androgen receptors [14], providing a biological rationale for examining this structure during the peri-pregnancy period. In our cohort, CP volume did not show a significant longitudinal change across the pre-pregnancy and postpartum MRI timepoints and was not retained in the final EDSS model. To our knowledge, this is the first study to longitudinally evaluate CP volume during the peri-pregnancy period in women with MS. Direct comparison with previous peri-pregnancy studies is therefore not possible. Longitudinal studies in MS outside the pregnancy setting suggest that the temporal course and clinical relevance of CP volume may vary. Klistorner et al. [16] found that CP volume remained stable over four years and was not associated with changes in EDSS. Bergsland et al. [36] also reported no significant increase in CP volume over five years and found no association between CP volume and subsequent disability progression. In contrast, CP pseudo-T2 was associated with progression [36]. When considered together with these longitudinal data, our findings suggest that volumetric CP measures alone may not capture all clinically relevant changes and that microstructural measures may provide additional information.

Although six patients experienced postpartum relapses, EDSS scores showed little overall change over follow-up. Previous pregnancy studies have similarly shown that increased relapse and radiological disease activity during the postpartum period are not always accompanied by a clear short-term increase in disability [4, 5, 37, 38]. Given the limited within-patient variation in EDSS, the observed volume–EDSS associations may partly reflect differences between patients rather than parallel within-patient changes over time. They should therefore not be interpreted as indicators of disability progression.

Several limitations should be considered. The retrospective, single-center design and small sample size limit statistical power and generalizability. The large number of candidate MRI-derived variables relative to the sample size also increases the risk of overfitting and model instability, which cannot be fully eliminated by forward stepwise selection. MRI scans were acquired as part of routine clinical care using different scanners and field strengths, and imaging intervals varied across patients. In addition, the absence of MRI data during gestation prevented direct characterization of structural changes occurring during pregnancy, including transient changes that may have resolved before postpartum imaging. Although longitudinal image-processing and harmonization methods were applied, residual technical effects cannot be excluded. The absence of healthy pregnant women and non-pregnant women with MS as control groups limits our ability to distinguish pregnancy-related physiological changes from MS-related structural changes and background longitudinal variability. Limited EDSS variability restricted the assessment of disability progression. The sample size also precluded robust evaluation of the effects of DMT exposure and treatment timing, breastfeeding duration, and relapse activity on regional brain volumes. Finally, hormonal measures and serum and CSF inflammatory biomarkers were unavailable, limiting mechanistic interpretation.

Future prospective studies should include standardized serial MRI assessments before conception, during gestation, and in the early postpartum period. The inclusion of healthy pregnant women and non-pregnant women with MS as control groups would help distinguish pregnancy-related physiological remodeling from MS-related structural changes. Larger cohorts would also allow the effects of DMT exposure, breastfeeding, and relapse activity to be examined more reliably. Combining regional volumetric measures with more sensitive clinical outcomes, hormonal and inflammatory biomarkers, and microstructural imaging may further clarify the clinical relevance of peri-pregnancy brain changes in MS. Such studies should also incorporate cognitive and neuropsychological assessments together with validated measures of mood, fatigue, and other relevant behavioral domains.

## Conclusion

This pilot study suggests that regional brain volumetric measures, particularly ventral diencephalon volume, may be associated with disability during the peri-pregnancy period in women with relapsing-remitting MS. The most prominent regional association was observed for the left ventral diencephalon. Choroid plexus volume did not show a significant longitudinal change and was not retained in the final EDSS model. These findings are exploratory and should be interpreted as hypothesis-generating. Larger prospective studies with standardized MRI protocols, predefined pre-pregnancy and postpartum imaging timepoints, appropriate control groups, integrated hormonal and inflammatory biomarkers, and sufficient power to evaluate DMT-related effects are needed to confirm these findings and clarify the biological mechanisms underlying peri-pregnancy neurostructural changes in MS. Nonetheless, this study represents an important step toward understanding the complex interplay between pregnancy, MS-related changes in brain volume, and their relationship with disability.

## Statements and Declarations

### Ethics Approval

This study was performed in accordance with the principles of the Declaration of Helsinki. The study protocol was approved by the Ethics Committee of Ondokuz Mayıs University (approval number: 2025/9).

### Consent to Participate

Written informed consent was obtained from all participants included in the study for the use of their clinical and MRI data for research purposes.

### Data Availability

The data that support the findings of this study are available from the corresponding author upon reasonable request.

### Author Contributions

M.T.S., C.T., K.B., and M.T. contributed to the conception and design of the study. M.T.S., S.K., D.A.M., and M.T. contributed to clinical data collection and curation. M.T.S. and S.K. selected and prepared the MRI data for neuroimaging analysis. K.B. and L.J.D. performed the MRI processing and neuroimaging analyses. C.T. performed the statistical analyses. M.T.S., K.B., and C.T. drafted the initial version of the manuscript. All authors contributed to data interpretation, critically revised the manuscript for important intellectual content, and approved the final version.

