## Supplemental Table for "Association Between Disability and Longitudinal Brain Volumetric Measures Across the Peri-Pregnancy Period in Women with Multiple Sclerosis: A Pilot Study"

**Supplementary Table.** Details of MRI scanners and acquisition parameters.

| Sequence | N | Magnetic Field Strength | Manufacturer | TE (ms) | TR (ms) | Flip angle (°) | Slice thickness (mm) | In-plane voxel size (mm) |
|---|---|---|---|---|---|---|---|---|
| T1 2D | 15 | 1.5T (n=11), 3.0T (n=4) | Philips (n=5), GE (n=5), Siemens (n=4), Hitachi (n=1) | 8.8–24.7 | 336–2898 | 69.0–160.0 | 4.0–5.0 | 0.41–1.00 |
| T1 3D | 23 | 1.5T (n=13), 3.0T (n=10) | Philips (n=16), GE (n=2), Siemens (n=5) | 2.9–11.1 | 7–2200 | 8.0–90.0 | 1.0–2.0 | 0.89–1.12 |
| FLAIR 2D | 15 | 1.5T (n=14), 3.0T (n=1) | Philips (n=9), Siemens (n=4), Hitachi (n=1), GE (n=1) | 78.0–140 | 3700–11000 | 90.0–180.0 | 4.0–5.0 | 0.47–1.00 |
| FLAIR 3D | 23 | 1.5T (n=10), 3.0T (n=13) | Philips (n=12), GE (n=6), Siemens (n=5) | 39.7–410 | 60–8000 | 20.0–120.0 | 1.0–3.0 | 0.41–1.12 |

MRI acquisition parameters are summarized separately for two-dimensional (2D) and three-dimensional (3D) T1-weighted and fluid-attenuated inversion recovery (FLAIR) sequences. N denotes the number of MRI scans available for each sequence type. Values for magnetic field strength and scanner manufacturer are presented as the number of MRI scans (n) for each sequence type. Continuous acquisition parameters are reported as the observed range across scans. TE, echo time; TR, repetition time; ms, milliseconds; mm, millimeters; T, Tesla; GE, GE Healthcare.
