## Supplemental Figure for "Association Between Disability and Longitudinal Brain Volumetric Measures Across the Peri-Pregnancy Period in Women with Multiple Sclerosis: A Pilot Study"

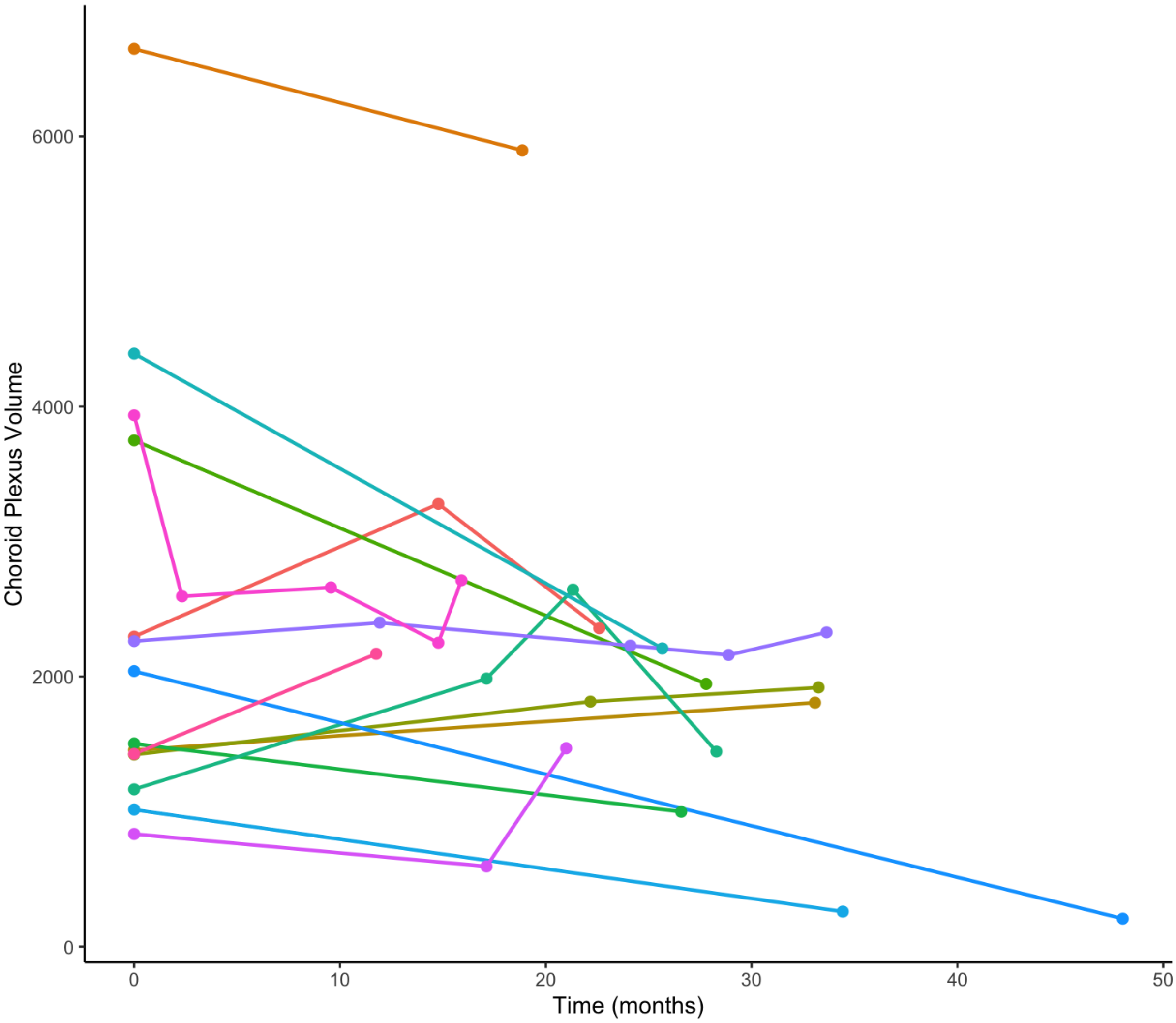


**Supplementary Figure.** Individual longitudinal trajectories of choroid plexus volume across the peri-pregnancy period. Each line represents a single participant across available MRI timepoints. Harmonized choroid plexus volumes are shown
